# Postpartum bleeding and shock in women giving birth with severe anaemia

**DOI:** 10.64898/2026.03.11.26348119

**Authors:** Raoul Mansukhani, Haleema Shakur-Still, Danielle Prowse, Amber Geer, Amy Brenner, Katharine Ker, Judith Lieber, Eni Balogun, Monica Arribas, Rizwana Chaudhri, Projestine Muganyizi, Oladapo Olayemi, Folasade Adenike Bello, Mwansa Ketty Lubeya, Bellington Vwalika, Ian Roberts

**Author notes:** Corresponding author: Raoul Mansukhani, Global Health Trials Group, London School of Hygiene & Tropical Medicine, London, WC1E 7HT.

## Abstract

**Background:** Each year, about 70,000 women die from bleeding after childbirth. Maternal anaemia increases the risk of postpartum bleeding. WHO recommends that treatment for postpartum haemorrhage should begin if there is blood loss ≥300 ml with abnormal haemodynamic signs (including elevated shock index) or blood loss ≥500 ml within 24 hours of birth. The prevalence of anaemia is highest in Sub-Saharan Africa and South Asia and these regions also have the highest maternal death rates. We examine the effect of severe anaemia (haemoglobin <70 g/L) compared to moderate anaemia (haemoglobin 70-99 g/L) on shock in 15066 women giving birth in Nigeria, Pakistan, Tanzania and Zambia.

**Methods:** We conducted a cohort analysis of data from the WOMAN-2 trial. We report mean and median estimated blood loss and the number of women with blood loss ≥1L, stratified by anaemia severity. We defined shock as post birth shock index >1.3. We define shock index as heart rate ÷ systolic blood pressure. We use multivariable logistic regression to calculate an odds ratio for the effect of severe anaemia on shock, adjusting for estimated blood loss, antepartum haemorrhage or placenta abnormalities, infection and hypertension.

**Results:** Mean (SD) estimated blood loss was 306 (177) ml in women with moderate anaemia compared to 339 (269) ml for severe anaemia. Median (IQR) estimated blood loss was 280 (200-350) ml for moderate anaemia versus 290 (200-360) ml for severe anaemia. Blood loss ≥1L occurred in 1.2% (156/12982) of women with moderate anaemia versus 2.9% (60/2076) with severe anaemia (OR=2.45 95%CI 1.81-3.31). Shock occurred in 0.8% (103/12,974) of women with moderate anaemia versus 2.3% (48/2,070) with severe anaemia. Severe anaemia tripled the odds of shock (OR=3.35, 95%CI 2.34-4.81). After adjusting for blood loss and other factors, severe anaemia more than doubled the odds of shock (aOR=2.39, 95%CI 1.59-3.58).

**Conclusions:** For any given blood loss, women with severe anaemia are more likely to become shocked. Preventing and treating anaemia in women of reproductive age must be a public health priority.

## Introduction

About 70,000 women die each year from bleeding after birth.^1^ Almost all of them are in low- and middle-income countries, mostly south Asia and sub-Saharan Africa.^1^ The World Health Organization (WHO) estimates that nearly half of women of reproductive age in South Asia and over one third in sub-Saharan Africa have anaemia.^2^ Compared to women with moderate anaemia, those with severe anaemia have twice the odds of a clinical postpartum haemorrhage (PPH) diagnosis.^3^

Based on a 2025 study analysing the relationship between blood loss, vital signs, and maternal death or severe morbidity, WHO introduced updated criteria for PPH treatment to prevent maternal mortality.^4,5^ The guidelines now recommend that treatment should begin if there is objectively measured blood loss ≥300 ml with any abnormal haemodynamic sign (including elevated shock index), or blood loss ≥500 ml, whichever occurs first within 24 hours of birth.^4^ Previous guidelines recommended treatment starting at blood loss ≥500 ml.^6^ An elevated shock index is often the result of severe bleeding but anaemia also has profound effects on the cardiovascular system.^7^ We use data from 15,066 women with moderate (haemoglobin 70-99 g/L) or severe anaemia (haemoglobin < 70 g/L) giving birth vaginally in hospitals in sub-Saharan Africa and South Asia to assess the effects of severe anaemia and blood loss on shock.^8^

## Methods

### Study design and data collection

We conducted a cohort study using data from the WOMAN-2 trial to examine the association between prebirth anaemia severity and both estimated blood loss and shock. The WOMAN-2 trial recruited women in 34 hospitals from four countries (Nigeria, Pakistan, Tanzania, and Zambia) where anaemia during pregnancy is commonly observed. Participants were recruited between 7 August 2019 and 19 September 2023 (Nigeria: 8 March 2020 – 19 September 2023; Pakistan: 7 August 2019 – 19 September 2023; Tanzania: 1 March 2022 – 19 September 2023; Zambia: 3 January 2020 – 16 September 2023). The methods and results are presented in detail elsewhere^8–11^. Briefly, we recruited women of any age with moderate or severe anaemia (haemoglobin <100 g/L) who were in active labour and about to give birth vaginally. We collected data on the woman’s health and pregnancy status and measured her prebirth blood pressure, heart rate and haemoglobin concentration at trial entry. Heart rate and blood pressure were measured using a Microlife Cradle VSA, and haemoglobin was measured in capillary blood using a portable HemoCue 201+.^12,13^ Women who were younger than 18 years without permission provided by a guardian, had a known tranexamic acid allergy, or developed postpartum haemorrhage before the umbilical cord was cut or clamped were excluded from the study. The lead obstetrician and trial team trained clinicians at each participating hospital to estimate blood loss in the 24 hours after birth or prior to hospital discharge. We used pictorial guides, with photographs of soiled pads and bed sheets, to aid the visual estimation of blood loss. Within 24 hours after birth or prior to hospital discharge, we recorded the lowest blood pressure and corresponding pulse rate, and any blood transfusions given.

The WOMAN-2 trial was approved by the London School of Hygiene & Tropical Medicine’s Ethics Committee (reference 15194) and the relevant ethics committees of all participating hospitals. We obtained informed consent from women if their physical and mental capacity allowed. If a woman was unable to give fully informed consent, information was provided to her level of capacity and her verbal agreement obtained in the presence of an impartial witness, with enrolment permitted on this basis. The opinion of the woman prevailed over that of accompanying persons. Fully informed consent was sought as soon as the woman regained capacity. The consent procedures are described in detail in the study protocol.^9^

### Statistical methods

To show how mean (95% CI) prebirth heart rate and systolic blood pressure varies with pre-birth haemoglobin, we plot point estimates with confidence intervals across 10 g/L haemoglobin categories.

We use box plots to show how median (IQR) estimated blood loss varies between women with moderate (haemoglobin 70-99 g/L) and severe anaemia (haemoglobin <70 g/L). We report mean (SD) and median (IQR) estimated blood loss for women with moderate and severe anaemia. We use t-tests (with Satterthwaite’s correction) to compare means across anaemia severity categories and Wilcoxon rank-sum tests to compare medians. We report means in addition to medians for blood loss stratified by anaemia severity because, although median blood loss may be similar, women with severe anaemia may be more likely to experience extreme blood loss values.^3^ To further examine how median blood loss varies by anaemia severity we categorised women as having moderate anaemia (prebirth haemoglobin of 70-99 g/L), severe anaemia (50-69g/L) or very severe anaemia (<50g/L). We use a Kruskal-Wallis test to assess the association between anaemia severity and median estimated blood loss.

We report an OR (95% CI) for the association between severe anaemia and blood loss ≥1L, a threshold previously used by WHO (2012) to define severe PPH and associated with increased risk of maternal mortality and need for transfusion. ^14,15^

Shock index is defined as heart rate ÷ systolic blood pressure.^16^ We define shock as shock index >1.3, a diagnostic threshold associated with increased mortality risk.^16^ We report prebirth shock rates by anaemia severity and used Fisher’s exact test to compare groups. To show how the risk of a postpartum shock varies with pre-birth haemoglobin, we plot point estimates with confidence intervals across 10 g/L haemoglobin categories. We report an OR (95% CI) for the association between severe anaemia vs moderate anaemia and shock using mixed effects logistic regression with random intercepts for hospital (nested within country) to account for clustering. We use multivariable mixed effects logistic regression to quantify the association between anaemia severity and shock, controlling for estimated blood loss. We repeat this analysis with additional adjustment for antepartum haemorrhage or placenta abnormalities, infection and hypertension. We select confounding variables which are putatively associated with both anaemia and high shock index showing our causal assumptions in a directed acyclic graph (appendix figure 1). We use a bar chart to show the proportion of women with shock across blood loss categories (<500, 500-999 and ≥1000 mL), with separate bars comparing women with moderate anaemia (red) and severe anaemia (pink).

**Figure 1a:**
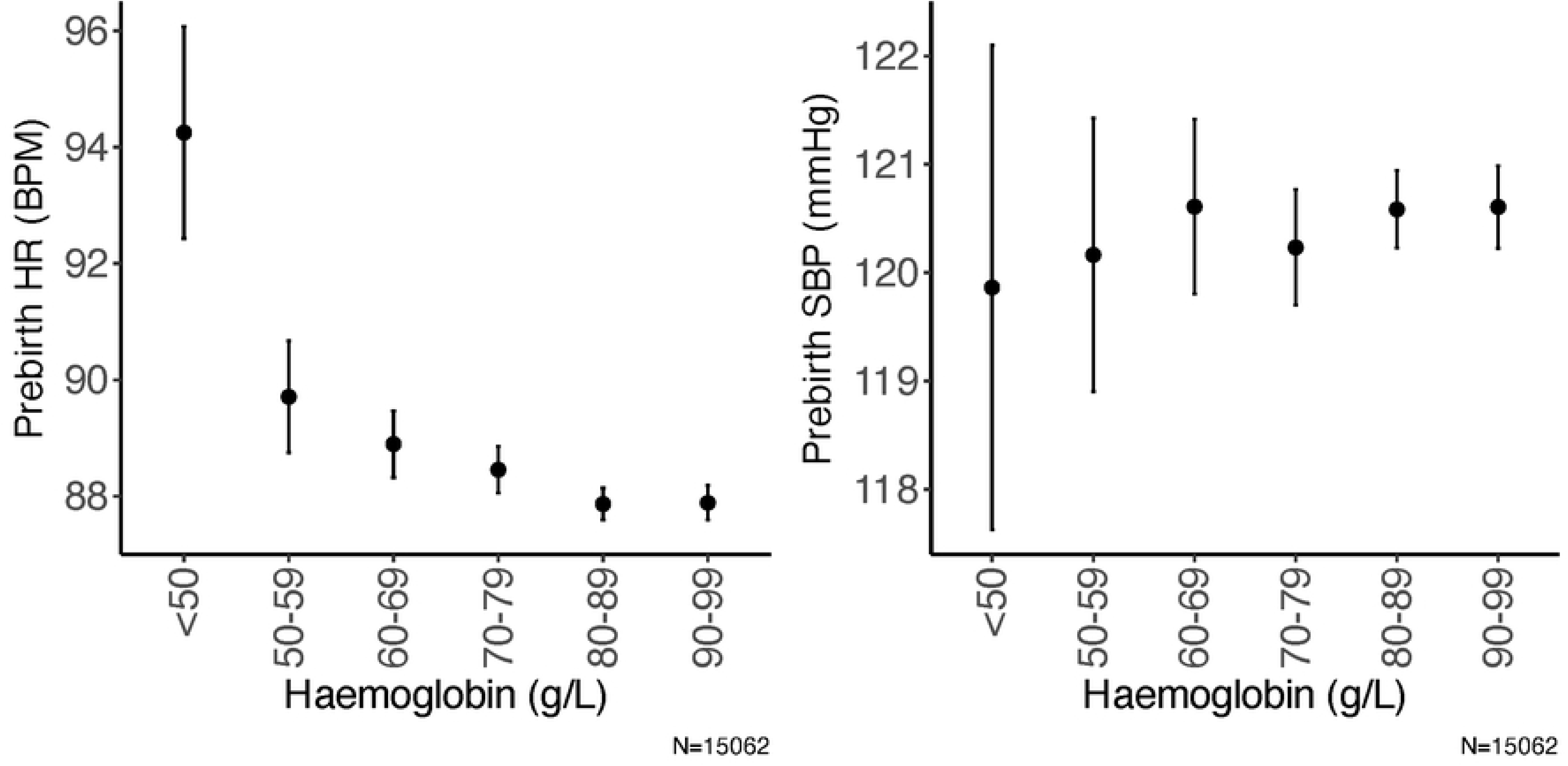
Prebirth haemoglobin vs prebirth heart rate (HR), N=15062. Figure 1b: prebirth haemoglobin vs prebirth systolic blood pressure (SBP), N=15062. Missing values: prebirth HR (4), prebirth SBP (4)

Estimated blood loss is subject to potential inaccuracies. To address this limitation and strengthen the robustness of our findings, we repeat key analyses replacing clinician estimated blood loss with the objective measure of calculated blood loss. Specifically, we: 1) repeat the adjusted analyses for the association between anaemia severity and shock, and 2) reproduce the bar chart showing the proportion of women with shock across blood loss categories. We obtain calculated blood loss from the product of estimated blood volume and relative peripartum change in haemoglobin. We obtain estimated blood volume (in litres) by multiplying maternal weight in kg by 0.085.^17^ We correct for the effect of transfusion on postpartum haemoglobin using methods described by Roubinian.^18^

We conduct complete case analyses rather than imputing missing values as we have a large sample size with minimal missingness.

## Results

The WOMAN-2 trial recruited 15,068 women from hospitals in Pakistan (73.2% of women), Tanzania (13.5%), Nigeria (8.8%), and Zambia (4.6%). Outcome data were obtained for 15,066 women. The mean (SD) age was 27 (6) years. Table 1 shows the characteristics of the participating women.

**Table 1:** Baseline characteristics of women with moderate or severe anaemia from the WOMAN-2 trial. Proportions may not sum to 100 % due to rounding. N=15066

| Variable | Number | % (SD) |
| --- | --- | --- |
| **Haemoglobin (g/L)** |  |  |
| Mean (SD) | 83 | (12) |
| Moderate anaemia | 12988 | 86.2 % |
| Severe anaemia | 2078 | 13.8 % |
| **Country** |  |  |
| Pakistan | 11024 | 73.2 % |
| Nigeria | 1326 | 8.8 % |
| Tanzania | 2029 | 13.5 % |
| Zambia | 687 | 4.6 % |
| **Age (Years)** |  |  |
| Mean (SD) | 27 | (6) |
| <20 | 895 | 5.9 % |
| 20-29 | 8936 | 59.3 % |
| 30-39 | 4829 | 32.1 % |
| 40+ | 406 | 2.7 % |
| **Placenta abnormalities** |  |  |
| Abruption | 425 | 2.8 % |
| Previa | 35 | 0.2 % |
| Abruption, Previa | 6 | 0.0 % |
| Accreta | 3 | 0.0 % |
| None | 14597 | 96.9 % |
| **Any antepartum haemorrhage** |  |  |
| Yes | 435 | 2.9 % |
| No | 14631 | 97.1 % |
| **APH or Placenta abnormalities** |  |  |
| Yes | 565 | 3.8 % |
| No | 14501 | 96.2 % |
| **Current infection status** |  |  |
| HIV | 283 | 1.9 % |
| Hepatitis | 157 | 1.0 % |
| Malaria | 19 | 0.1 % |
| Syphilis | 15 | 0.1 % |
| Other | 96 | 0.6 % |
| None | 14515 | 96.3 % |
| **Hypertensive disease** |  |  |
| Eclampsia | 41 | 0.3 % |
| Pre-eclampsia | 321 | 2.1 % |
| Pre-existing hypertension | 55 | 0.4 % |
| Pregnancy-induced hypertension | 734 | 4.9 % |
| None | 13920 | 92.4 % |
| **Estimated blood loss (ml)** |  |  |
| Mean (SD) | 310 | (193) |
| <500 | 13983 | 92.8 % |
| 500-999 | 859 | 5.7 % |
| 1000+ | 216 | 1.4 % |
| Missing | 8 | 0.1 % |

Figures 1a and 1b show how mean (95% CI) prebirth heart rate and systolic blood pressure vary with baseline haemoglobin. Mean heart rate declined as haemoglobin increased, whereas mean systolic blood pressure showed only small variation. Prebirth shock (shock index >1.3) occurred in 0.6% (12/2,078) of women with severe anaemia compared to 0.1% (9/12,984) with moderate anaemia (p<0.001).

Figure 2 shows median (IQR) estimated blood loss in women with moderate vs severe anaemia. Mean (SD) estimated blood loss was higher in women with severe anaemia compared to those with moderate anaemia (339 (269) vs 306 (177) ml; p<0.001).

**Figure 2:**
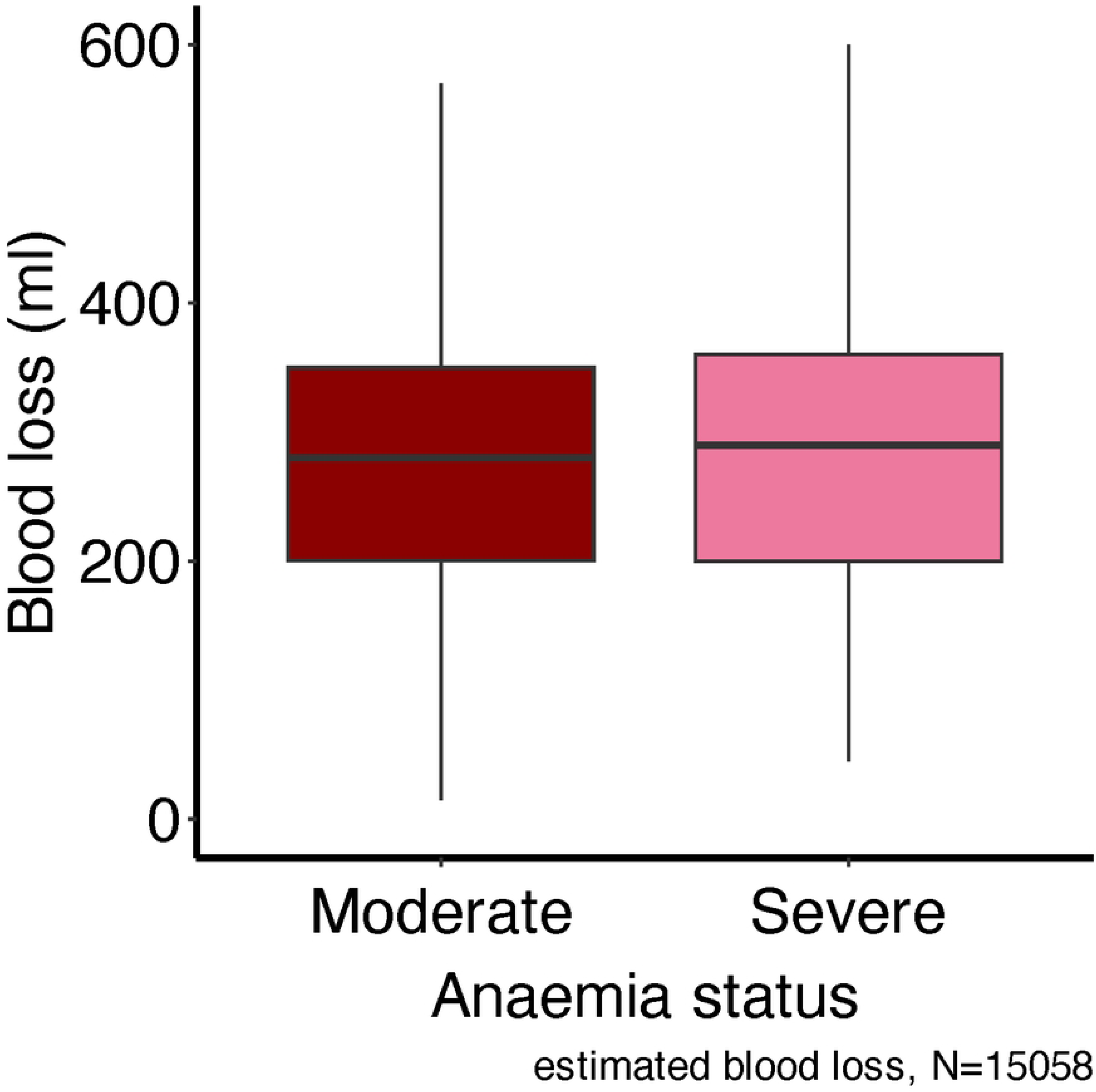
Estimated blood loss in all women. Moderate anaemia (70-99 g/L), severe anaemia (<70 g/L). WOMAN-2 trial data. N=15058. Missing values: estimated blood loss (8).

Median (IQR) estimated blood loss was (280 (200-350) ml vs 290 (200-360) ml; p=0.021) for moderate vs severe anaemia respectively. Using a three-category classification, median blood loss was 280 ml (IQR 200-350) for haemoglobin 70-99 g/L, 290 ml (IQR 200-350) for haemoglobin 50-69 g/L, and 300 ml (IQR 200-400) for haemoglobin <50 g/L.

Estimated blood loss of ≥1 L occurred in 1.2% (156/12,982) of women with moderate anaemia and 2.9% (60/2076) with severe anaemia. Women with severe anaemia had more than twice the odds (OR=2.45 95%CI 1.81-3.31) of having an estimated blood loss ≥1 L.

Figure 3 and table 2 show the relationship between prebirth haemoglobin and risk of postpartum shock. Risk increased (p<0.001) as haemoglobin dropped: 0.8% (70-99 g/L), 1.8% (50-69 g/L), and 6.8% (<50 g/L).

**Figure 3:**
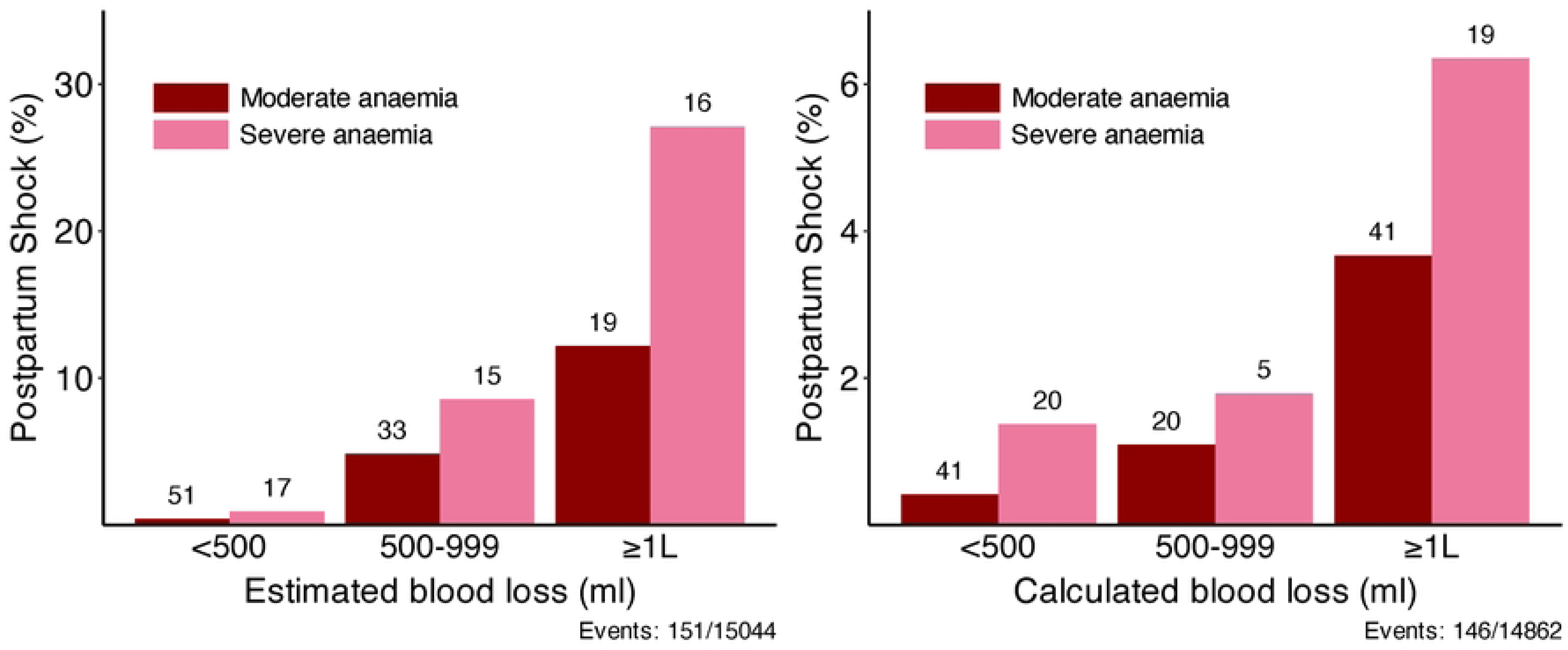
Prebirth haemoglobin versus risk of shock. Shock is defined as shock index >1.3. WOMAN-2 trial data. N=15046. Missing values: shock (20)

**Table 2:** Anaemia severity versus both median estimated blood loss and risk of shock. Shock is defined as shock index >1.3. WOMAN-2 trial data. Missing values: Estimated blood loss (8), shock (20).

| Variable | Moderate<br>70-99 g/L<br>(N=12988) | Severe<br>50-69 g/L<br>(N=1859) | Very severe<br><50 g/L<br>(N=219) | P value |
| --- | --- | --- | --- | --- |
| Estimated blood loss (ml) | 280 (200-350) | 290 (200-350) | 300 (200-400) | 0.046 |
| Shock (%) | 0.8% (103/12975) | 1.8% (33/1852) | 6.8% (15/219) | <0.001 |

Table 3 shows 0.8% (103/12,974) of women with moderate anaemia and 2.3% (48/2,070) with severe anaemia had postpartum shock. Women with severe anaemia had three times the odds of shock compared to those with moderate anaemia (OR=3.35, 95%CI 2.34-4.81). After controlling for estimated blood loss, the odds ratio decreased to 2.63 (95%CI 1.79-3.87). With further adjustment for antepartum haemorrhage or placental abnormalities, infection and hypertension, severe anaemia remained associated with more than twice the odds of shock (aOR=2.39, 95%CI 1.59-3.58). Results were similar when substituting calculated blood loss for estimated blood loss (aOR=2.26, 95%CI 1.52-3.36). Full details of our adjusted models are presented in tables 4a and 4b. Figure 4 shows that at every level of blood loss, women with severe anaemia had higher rates of shock than those with moderate anaemia. This result is consistent whether blood loss is estimated by a clinician or based on peripartum change in haemoglobin.

**Table 3:** The association between prebirth severe anaemia and shock. OR**†** adjusted for estimated blood loss. Adj. OR is adjusted odds ratio, controlling for estimated blood loss, antepartum haemorrhage or placenta abnormalities, infection and hypertension. Unadjusted and adjusted ORs include a random effect adjustment for hospital nested within country. WOMAN-2 trial data. (N=15044). Shock is defined as shock index >1.3. **◊** Results adjusted for calculated blood loss (N=14862). Missing values: Estimated blood loss (8), shock (20), Calculated blood loss (203)

| Variable | Moderate | Severe | OR | OR † | Adj. OR |
| --- | --- | --- | --- | --- | --- |
| Shock | 0.8% (103/12974) | 2.3% (48/2070) | 3.35 (2.34-4.81; p<0.001) | 2.63 (1.79-3.87; p<0.001) | 2.39 (1.59-3.58; p<0.001) |
| Shock ♦ |  |  |  | 2.66 (1.82-3.89; p<0.001) | 2.26 (1.52-3.36; p<0.001) |

**Table 4a.** Crude and adjusted odds ratios for risk factors of postpartum shock in women with moderate or severe anaemia. Shock is defined as shock index >1.3. Adjustment is made for estimated blood loss, antepartum haemorrhage or placenta abnormalities, infection, and hypertension. Unadjusted and adjusted ORs include a random effect adjustment for hospital nested within country. WOMAN-2 trial data. N=15044. Missing values: Shock (20), estimated blood loss (8)

| Variable | Total | Number of women<br>with shock (%) | OR (95% CI) | aOR (95% CI) |
| --- | --- | --- | --- | --- |
| **Severe anaemia** | 2070 | 48 (2.3) | 3.35 (2.34-4.81) | 2.39 (1.59-3.58) |
| **Est. blood loss** |  |  |  |  |
| <500 | 13972 | 68 (0.5) | 1.00 | 1.00 |
| 500-999 | 857 | 48 (5.6) | 13.34 (9.01-19.75) | 12.65 (8.46-18.90) |
| ≥1L | 215 | 35 (16.3) | 57.03 (35.08-92.73) | 48.37 (28.63-81.73) |
| **APH / abnormal placenta** |  |  |  |  |
| No | 14481 | 124 (0.9) | 1.00 | 1.00 |
| Yes | 563 | 27 (4.8) | 5.54 (3.55-8.63) | 1.50 (0.87-2.58) |
| **Current infection** |  |  |  |  |
| No | 14494 | 134 (0.9) | 1.00 | 1.00 |
| Yes | 550 | 17 (3.1) | 1.82 (1.02-3.24) | 1.78 (0.98-3.21) |
| **Hypertensive disease** |  |  |  |  |
| No | 13900 | 131 (0.9) | 1.00 | 1.00 |
| Yes | 1144 | 20 (1.7) | 1.64 (1.01-2.66) | 0.80 (0.47-1.37) |

**Table 4b.** Crude and adjusted odds ratios for risk factors of postpartum shock in women with moderate or severe anaemia. Shock is defined as shock index >1.3. Adjustment is made for calculated blood loss, antepartum haemorrhage or placenta abnormalities, infection and hypertension. Unadjusted and adjusted ORs include a random effect adjustment for hospital nested within country. WOMAN-2 trial data. N= 14862. Missing values: shock (20), calculated blood loss (203)

| Variable | Total | Number of women<br>with shock (%) | OR (95% CI) | aOR (95% CI) |
| --- | --- | --- | --- | --- |
| **Severe anaemia** | 2037 | 44 (2.2) | 3.08 (2.13-4.47) | 2.26 (1.52-3.36) |
| **Calc. blood loss** |  |  |  |  |
| <500 | 11339 | 61 (0.5) | 1.00 | 1.00 |
| 500-999 | 2107 | 25 (1.2) | 2.17 (1.35-3.48) | 2.14 (1.33-3.43) |
| ≥1L | 1416 | 60 (4.2) | 7.46 (5.14-10.81) | 6.24 (4.26-9.13) |
| **APH / abnormal placenta** |  |  |  |  |
| No | 14316 | 122 (0.9) | 1.00 | 1.00 |
| Yes | 546 | 24 (4.4) | 5.04 (3.16-8.02) | 2.64 (1.59-4.40) |
| **Current infection** |  |  |  |  |
| No | 14320 | 129 (0.9) | 1.00 | 1.00 |
| Yes | 542 | 17 (3.1) | 1.87 (1.05-3.34) | 1.70 (0.94-3.08) |
| **Hypertensive disease** |  |  |  |  |
| No | 13734 | 129 (0.9) | 1.00 | 1.00 |
| Yes | 1128 | 17 (1.5) | 1.41 (0.84-2.36) | 0.91 (0.53-1.56) |

**Figure 4:**
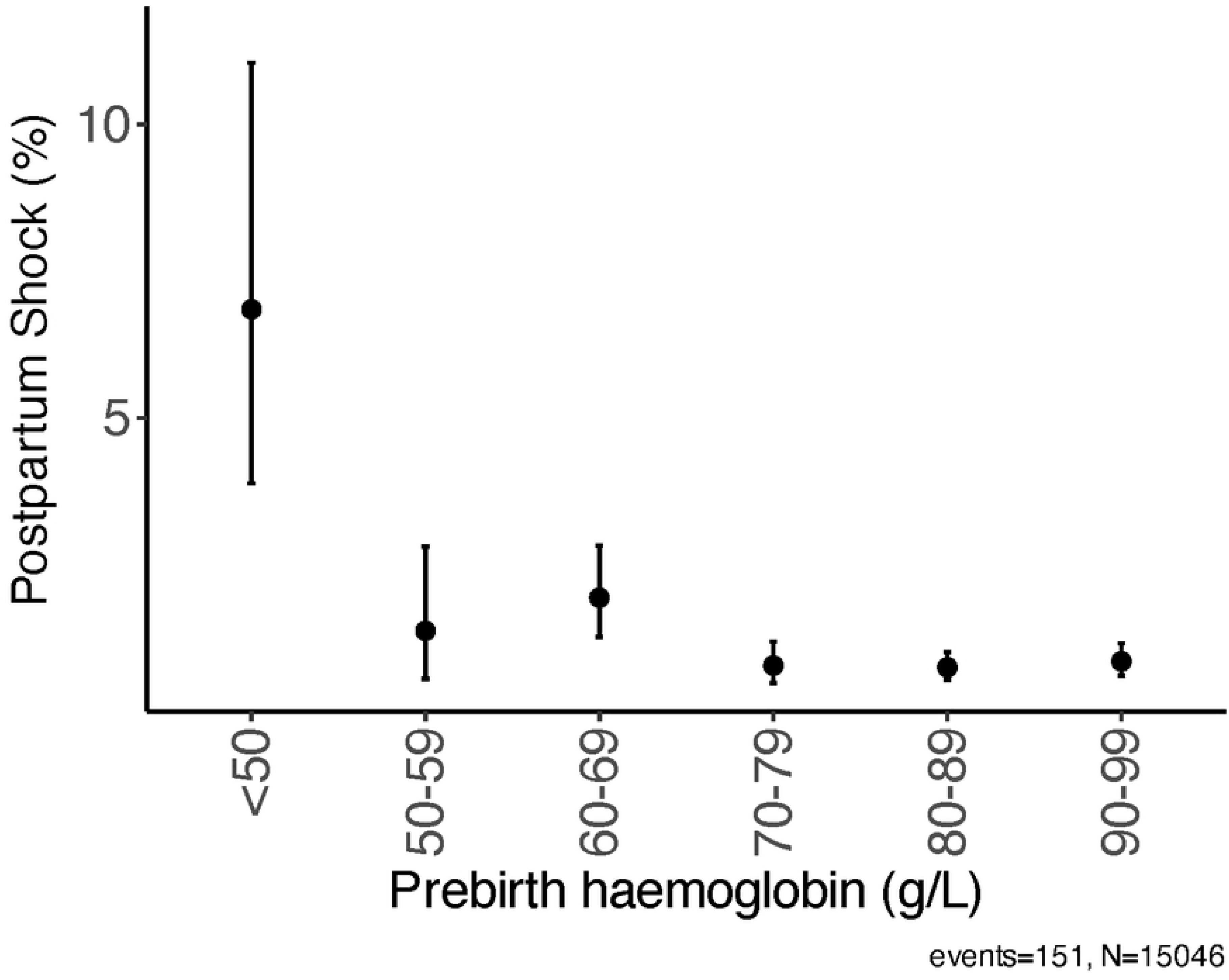
Risk of shock by estimated blood loss category. N=15044. **Figure 4b**: Risk of shock by calculated blood loss category. N=14862. Estimated blood loss is obtained from visual inspection of soiled sheets and pads. Calculated blood loss is obtained by proportional peripartum change in haemoglobin and maternal weight. Shock is defined as shock index >1.3. The number of women with shock is shown above each bar. WOMAN-2 trial data. Missing values: shock (20), estimated blood loss (8), calculated blood loss (203)

Our data had 8 (0.1%) missing values for estimated blood loss. We were unable to calculate shock for 20 (0.1%) women due to either missing heart rate or systolic blood pressure measurements and we were unable to obtain calculated blood loss for 203 (1.3%) women due to missing postpartum haemoglobin measurements.

## Discussion

We show that for any given blood loss, women with severe anaemia are more likely to become shocked. After controlling for blood loss, women with severe anaemia had more than twice the odds of shock index > 1.3.

Women with severe anaemia were more likely to experience extreme blood loss, as demonstrated by higher rates of blood loss ≥ 1L. Mean blood loss was higher in women with severe anaemia compared to those with moderate anaemia, although median values were similar.

We found an inverse relationship between haemoglobin and prebirth heart rate, with stable blood pressure. More women with severe anaemia already had shock before giving birth compared to those with moderate anaemia.

To the best of our knowledge, ours is the first study to show that prepartum severe anaemia is an independent risk factor for postpartum shock in women giving birth. We collected data for women giving birth in sub-Saharan Africa and South Asia, settings with the highest maternal death rates world-wide. A recent systematic review, including all population-based cohort studies published between January 1960 and November 2024 reporting risk factors for PPH, confirmed the association between anaemia and postpartum haemorrhage.^19^ However, our study collected data on blood loss, postpartum shock and other maternal factors, with almost no loss to follow-up and minimal missing data.

We acknowledge our study has some weaknesses. While clinicians at each site were trained in estimating blood loss from soiled sheets and pads there will inevitably be some mismeasurement. When we repeated our analyses with calculated blood loss, based on peripartum change in haemoglobin, our results were almost identical. While haemoglobin was measured using capillary haemoglobinometry (HemoCue) which is suitable for screening purposes, there is some discrepancy with laboratory gold standard.^20^ We would expect any potential random error in haemoglobin measurement to attenuate our results. Although we adjusted for confounding factors, there will be some confounding factors that we were unable to control for. As all participants had moderate or severe anaemia, a comparison of the effects of severe anaemia with a cohort of women with no or mild anaemia was not possible. Women were enrolled from hospitals in Pakistan, Nigeria, Zambia and Tanzania, but most were recruited in Pakistan. Nevertheless, we see no biological reason why the association between severe anaemia and blood loss and shock that we observed would not be generalisable to women giving birth out of hospital or in other countries.

Shock is a state of reduced end organ oxygenation arising when tissue oxygen demand exceeds supply causing an oxygen debt.^21^ The amount of oxygen supplied to the tissues depends on the blood flow and the arterial oxygen content.^22^ Severe bleeding causes shock by reducing blood flow. However, because the arterial oxygen content depends on the haemoglobin concentration, oxygen supply is also reduced in severe anaemia and women with severe anaemia can develop organ failure from only modest blood loss. This has implications for management. At very low haemoglobin concentrations, sympathetic activation leads to reduced renal blood flow with salt and water retention causing oedema and cardiac failure.^23,24^ Women with anaemic cardiac failure are often hypervolemic and may be harmed by excessive intravenous fluids. If a high shock index in women with severe anaemia is mistaken for haemorrhagic hypovolaemia and treated with vigorous fluid resuscitation, this could cause pulmonary oedema. Transfusion Associated Cardiac Overload (TACO) is the leading cause of transfusion associated death in high income countries and is likely to be concern elsewhere.^25^ More research is needed to find the best ways to manage women giving birth with severe anaemia.

Women with severe anaemia are more likely to lose ≥1L of blood and are at increased risk of shock after giving birth. The WHO goal of halving the prevalence of anaemia in women of reproductive age by 2025 was not met and the target date has been reset to 2030^26^. Our data underscore the global health importance of preventing and treating anaemia before birth.

## Data Availability

After publication of primary and secondary analyses detailed in the statistical analysis plan, individual de-identified patient data, including the data dictionary, will be made available via our data sharing portal, The Free Bank of Injury and Emergency Research Data (FreeBIRD), indefinitely to allow maximum use of the data to improve patient care and advance medical knowledge. The trial protocol and statistical analysis plan are freely available online. https://freebird.lshtm.ac.uk

## Author contributions

RM and IR planned the analyses. RM and IR drafted the manuscript with input from all authors. RM did the statistical analyses, interpreted the data, and generated the figures. HS-S and IR were the chief investigators of the WOMAN-2 trial. RC was lead coordinating investigator in Pakistan. FAB and OO were lead coordinating investigators in Nigeria. PM was lead coordinating investigator in Tanzania. MKL and BV were responsible for data collection in Zambia. KK, AB, MA and JL interpreted the data. EB was responsible for overall study management. AG and DP managed the data collection. RM, DP, EB, AB, MA and AG had direct access to the data and verified the data reported in the manuscript. All authors were involved in the development of the original manuscript, read and approved the final version submitted.

## Data availability

After publication of primary and secondary analyses detailed in the statistical analysis plan, individual de-identified patient data, including the data dictionary, will be made available via our data sharing portal, The Free Bank of Injury and Emergency Research Data (FreeBIRD), indefinitely to allow maximum use of the data to improve patient care and advance medical knowledge. The trial protocol and statistical analysis plan are freely available online.

## Acknowledgements

We are deeply indebted to Julio Gil Onandia and Collette Barrow for their incredible support in the preparation of this manuscript. We would like to thank all the study sites and staff who made the WOMAN-2 trial happen and most importantly, the participating women, without whom the study would not have been possible.

